# Reproducibility and Robustness of Localized Mortality Prediction

**DOI:** 10.1101/2024.06.04.24308417

**Authors:** Evelyn Nitch-Griffin, Amy Peterson, Yara Skaf, Jason Cory Brunson

## Abstract

**Background:** While localized modeling—the use of predictive models to perform the adaptation step in case-based reasoning—has been evaluated in several experimental settings, its reported successes have infrequently been independently and externally validated.

**Objective:** We aimed to extend and validate an experimental study of mortality prediction in a critical care population and to assess the importance of several methodological factors to predictive performance.

**Methods:** We reproduced the workflow of Lee, Maslove, and Dubin (2015) using an updated database. We evaluated performance as area under the receiver operating characteristic curve and under the precision–recall curve and calibration as weakness of evidence in Hosmer– Lemeshow tests. We compared the effects of several modeling choices, including how relevance is quantified, and how relevance cohorts are retrieved, and the choice of model. We compared ours to previous results and used linear regression to quantify the role of each modeling choice on performance.

**Results:** Overall performance and its relationship to cohort size validated previous results. These relationships varied by model family as expected, though we observed no advantage of decision trees over a model-free approach and poor performance by random forests. An alternate choice of unlearned similarity measure yielded marginal and inconsistent performance differences. Denominating cohorts by similarity threshold rather than by cardinality yielded marginal but consistent performance losses. A temporal validation exercise enabled by a change of information system before the recent upgrade corroborated performance estimates from cross-validation. In all, cohort denomination mattered more to performance than any other methodological choice.

**Discussion:** The greater impact of retrieval than of adaptation suggests a weakness with the strategy of localized modeling. Additional research to deconstruct the varieties of this approach and quantify the relative benefits of its components is needed to resolve this question.

## 1 Introduction

Case-based reasoning (CBR) emerged in the early years of artificial intelligence and saw extensive medical and clinical applications (Bichindaritz and Marling 2006). The technique has been improved through advances in its constituent stages, notably retrieval, applied to novel tasks such as surveillance and refinement of clinical guidelines, and integrated into a variety of health care systems (Bichindaritz and Montani 2011). It has also been extended to new, often hybrid techniques that build on elements of CBR. One of these techniques we call *localized modeling*. A localized model is any quantitative model fitted to a subset of data determined to be of greatest relevance to a specific case. Despite their reported merits, localized models have not, to our knowledge, been widely or routinely adopted in practice or generated sustained interest from the research community, nor have their successes been independently and externally validated.

Several dimensions of external validation are of practical interest: Target population, data collection, data pre-processing, model implementation, and model specification may all be required to validate a model, and certainly to put it into use, even if the domain and use case remain the same (Collins et al. 2014). The objective of this study is to assess the relative importance of these factors to the validation of an innovative application of localized models to mortality prediction in critical care settings.

### 1.1 Original Contributions

In this study we extend and validate a study by Lee, Maslove, and Dubin (2015) in three respects that are crucial to ensuring the generalizability of localized prediction modeling.

First, we use data from the release MIMIC-III (Johnson et al. 2016), which is much expanded from MIMIC-II. The pre-defined variables used by Lee, Maslove, and Dubin (2015) required manual redefinition for use with MIMIC-III, so that our study can be viewed as a robustness test of these preprocessing steps. The update presents an additional opportunity for robustness-testing: Between MIMIC-II and MIMIC-III, the hospital switched clinical information systems (CIS) from CareVue to MetaVision. The database from which records were obtained, reflecting the CIS in service at the time, is included as the database variable. We use this natural temporal partition of the stays to perform a temporal validation. This provides a more realistic evaluation of localized modeling as it would be applied in practice (Major, Jethani, and Aphinyanaphongs 2020).

Second, we compare the modified cosine similarity measure proposed by Lee, Maslove, and Dubin (2015), to an alternative: the Gower distance, a generalized distance measure for heterogeneous data. We use this measure as a no less natural or appropriate counterpart to cosine similarity. Both measures depend heavily on domain knowledge for variable selection. (Unreported results using domain-agnostic measures were significantly worse.)

Third, we compare model performance using two methods of cohort retrieval: *combinatorial* (in the sense of Goyal, Lifshits, and Schütze (2008)), obtaining cohorts of approximately equal cardinality, as used by Lee, Maslove, and Dubin (2015), and *statistical*, obtaining cohorts using a fixed similarity threshold, as distinguished by Park, Kim, and Chun (2006). While both methods have been used in practice, we are not aware of any comparisons of their performance or other utility since Park, Kim, and Chun (2006). The choice manifests the trade-off between relevance and statistical power, and the lack of published evidence to inform it is conspicuous.

To reduce computational cost and, we believe, present a more realistic use case, we make one fundamental change to the approach of Lee, Maslove, and Dubin (2015): We shift focus from one study population comprising all ICU admissions to several study populations comprising each the admissions to each ICU. This means that our study populations are more homogeneous in crucial respects, and that we no longer use the unit of admission as a predictor. We did not begin the project with strong expectations of how this might impact results, so our validation will be as much qualitative as quantitative.

### 1.2 Organization

We provide background on localized models, review validation approaches, and state our goals in Section 2. In Section 3 we describe our data source, our pre-processing steps, the experiments we conducted, the analyses we conducted, and the software we used. We survey our results in Section 4, roughly in order from most to least comparable to Lee, Maslove, and Dubin (2015). Finally, we contextualize our results and draw general lessons for localized modeling in Section 5, as well as note several limitations of our study and state our conclusions.

## 2 Background

### 2.1 Localized Models

In the forthcoming review, we proposed the name “localized modeling” for a family of techniques that involve fitting (usually predictive) models on cohorts of training data matched by relevance to testing data, rather than on the entire set of training data. These techniques are special kinds of case-based reasoning (CBR) in which the adaptive step, which obtains a candidate solution for a testing case from the known solutions to a set of matched training cases, is performed automatically by a quantitative model. This model can be from any statistical family, and previous studies have used logistic regression (Kenney Ng et al. 2015; Lee, Maslove, and Dubin 2015; Lee 2017; N. Wang et al. 2019), survival models (Mariuzzi et al. 1997; Lowsky et al. 2013), random forests (Lee 2017; N. Wang et al. 2019), fuzzy rules (Kasabov and Hu 2010; Liang, Hu, and Kasabov 2015; Verma et al. 2015), and other families (Vilhena et al. 2016; Ma et al. 2020; Y. Wang et al. 2020; K. Ng et al. 2021). Most studies also tune the sizes of the matched cohorts, which can range in cardinality from 1 to the size of the training data but may also be determined by another threshold, often a similarity threshold. Viewed as a family of models parameterized by cohort size, the localized models interpolate between nearest-neighbors prediction in which a prediction is obtained as an average or vote from the matched cohort, and the global fit of the model family.

### 2.2 External Validation

One dynamic at play between large health data and advanced predictive models is the struggle to keep method development apace with data availability: Health-related data sets obtained for secondary research use from databases of insurance claims, electronic health records, patient registries, and other sources are much larger and heterogeneous than conventional analysis techniques were designed for. Improvements in performance, memory, and parallelizability have addressed increased size, while new statistical transformations and models have accommodated increased diversity.

There is also an inverse dynamic at work that poses its own challenges: Data availability has struggled to keep up with the need to reproduce results in new settings. These data sets may be commonplace in the habitats for which they were designed, but their secondary use requires additional preparation that can be prohibitively resource-intensive. The result is that few or no research-ready data sets may be sufficiently similar to a previously studied data set—in terms of the sub-populations or risk factors being studied—to provide a suitable validation.

Predictive models may be validated *internally*, usually meaning on a subset of the data partitioned out before training and optimization, or *externally*, on data sets obtained under different circumstances from those used to train the model. The nature of these circumstances varies across contexts, particularly those that use large, heterogeneous, and variable-quality and -completeness data sets. Each of these qualities expands the range of preprocessing and analysis choices and therefore to the possibilities for external validation. External validation is more difficult than internal validation but all the more necessary to ensure that results generalize. The update from MIMIC-II to MIMIC-III (Section 3), which incorporated data collected after a transition between different EHR systems that affected several aspects of data collection and storage, provides a natural setting to test the external validity of novel methods originally demonstrated on MIMIC-II.

### 2.3 Objectives

From this perspective, we focus on the following questions:

- Do the results of Lee, Maslove, and Dubin (2015) obtain for an updated database and re-processed analytic data set?
- To what extent does the method of cohort retrieval matter, and what methods yield better-performing models?
- Are performance estimates based on temporal validation consistent with those based on folded cross-validation?

## 3 Materials and Methods

### 3.1 MIMIC-III

MIMIC-III (“Medical Information Mart for Intensive Care”) is a comprehensive open-access database compiled from the administrative and clinical records of over 40,000 patients who stayed within the intensive care units at Beth Israel Deaconess Medical Center between 2001 and 2012. It is maintained by the MIT Laboratory for Computational Physiology and collaborating research groups and has been widely used for education and research (Goldberger et al. 2000; Johnson et al. 2016). The exceptionally rich database is compiled from admission records, bedside monitoring devices, patient charts, laboratory tests, prescriptions, billing claims, clinical notes, and comprehensive death records and includes information such as demographics, bedside vital sign measurements, test results, procedures, medications, caregiver notes, imaging reports, and in-hospital and out-of-hospital mortality.

Researchers can access the complete database after completing a short online course and submitting a project description. The MIMIC-III maintainers provide a set of scripts to derive tables and variables commonly used in studies, such as severity scores, at the mimic-code repository (Johnson, Pollard, and Mark 2016). The repository is continually updated with code from the maintainers as well as from the international community of users. This shared resource by an active community increases the efficiency, reproducibility, and transparency of research on MIMIC-III. We have access to MIMIC-III through a predictive modeling and network analysis project initiated as part of a research experience for undergraduates, through which we have also contributed to the mimic-code repository. Data was taken from patients admitted to the medical ICU (MICU), surgical ICU (SICU), coronary care unit (CCU), cardiac surgery recovery unit (CSRU), and the trauma surgical ICU (TSICU). Since MIMIC-III v1.4 contains public, de-identified patient data, the project did not require review by an internal review board.

### 3.2 Patient Data Extraction

MIMIC-III contains subject IDs for individuals, hospital admission IDs for each distinct admission to the hospital, and ICU stay IDs for each distinct ICU admission. A patient could be discharged from the ICU and remain at the hospital, only to be readmitted to the ICU at a later date. Such a patient would have only one hospital admission ID but multiple ICU stay IDs. In this study, each ICU stay ID was treated as a single case, or data point. As explained in Lee, Maslove, and Dubin (2015), this allows our analysis to rely on similar cases rather than similar patients, as a CDS tool would in practice. For simplicity, we refer to ICU stays as “patients” in this article.

We extracted and calculated predictor variables to as closely as possible match those of Lee, Maslove, and Dubin (2015). Only patient data gathered in the first 24 hours after admission to the ICU was considered. For each ICU admission, the following sets of predictors and their summaries were computed:

- the maximum and minimum values for non-overlapping 6-hour periods of the following: heart rate, mean blood pressure, systolic blood pressure, SpO2, and body temperature;
- The minimum and maximum values during the first 24 hours of the following: hematocrit, white blood cell count, serum glucose, serum HCO3, serum potassium, serum sodium, blood urea nitrogen, and serum creatinine;
- the following categorical variables: admission type (elective, urgent, emergency), gender, ICU service type (MICU, SICU, CCU, CSRU, TSICU), primary ICD-9 code, the receipt of vasopressor therapy during the first 24 hours in the ICU (binary), and the use of mechanical ventilation or Continuous Positive Airway Pressure during the first 24 hours in the ICU (binary);
- age, minimum (worst) Glasgow Coma Scale, and total urinary output from each non-overlapping 6-hour period during the first 24 hours in the ICU; and
- the SAPS-II score and mortality likelihood, which required the extraction of bilirubin, PaO_2_, FiO_2_, and whether or not the patient was diagnosed with HIV, metastatic cancer, or hematologic malignancy, which were not used by Lee, Maslove, and Dubin (2015).

We extracted post-discharge 30-day mortality as our primary outcome and 30-day readmission as a secondary outcome.

Only patients with complete data were included in the analysis, meaning that all records missing data for any of the above variables were dropped. The only exception was non-overlapping 6-hour period total urinary output: If a patient had no data at all, they were dropped, but if they had no data for only one to three 6-hour periods, urinary output was assumed to be zero for that period.

#### 3.2.1 Changes from MIMIC-II

MIMIC-III contains all of the data in MIMIC-II, plus additional data obtained during 2008–2012. In 2008, the medical center switched from its previous system Carevue to a new data management system called Metavision. This involved restructuring certain aspects of how the data was stored and named. In particular, each variable was given a new item ID. Since there is no master list that provides a one-to-one crosswalk between Metavision and Carevue values, item IDs were determined by examining labels, consulting the code repository, and examining the distributions of relevant items in the case of uncertainty.

MIMIC-II came equipped with several calculated values, including SAPS-II and SOFA scores, first-day labs, and more, that were not included with MIMIC-III. Instead, PostgreSQL code to calculate these values is provided separately through the MIMIC code repository (Johnson, Pollard, and Mark 2016). We recapitulated these calculations in R rather than use the provided code, and we calculated SAPS-II but not SOFA. We dropped spontaneous respiratory rate from the analysis due to it being an estimated value at the discretion of the attending clinician.

### 3.3 Experiments

We adapted the model families of Lee, Maslove, and Dubin (2015): raw proportion (equivalent to nearest neighbors prediction under combinatorial retrieval), logistic regression (LR), and decision trees (DT), with their choices of hyperparameters. We considered also using random forests, following Lee (2017), but these were much more computationally expensive to fit and, when the number and maximum depth of the trees was reduced from the original, turned out to perform much worse than all three other model families.

For each critical care unit, and for each choice of similarity measure, retrieval denomination, and model family, we fit and evaluated models across a range of cohort sizes, using both folded and temporal validation. Combinatorial cohorts started at cardinality 10 and then ranged from 100 to the size of each training set at increments of 100, while statistical cohorts ranged in threshold across the theoretical limits of the similarity measure at increments of 0.01.

### 3.4 Analysis

We fitted and evaluated models in three phases:

1. Preprocess data from the MIMIC tables into rectangular case-by-variable data frames.
2. Partition each data frame from phase 1 into 10 strata for folded cross-validation and by database (CareVue or MetaVision) for temporal validation. For each pair of parts, generate matrices of similarity measures between training and testing data and of similarity rankings of training data from testing data.
3. Use the matrices from phase 2 to retrieve cohorts, fit models to these cohorts, generate predictions for their testing data, and summarize the performance of these predictions.

For purposes of this study, relevance is determined by a similarity or distance measure. In addition to the similarity measure used to match cases and the model family fitted to the similarity cohort, we parameterized over our localized models by the *denomination* and the *size* of the similarity cohorts. The *denomination* refers to whether cohorts are defined by a similarity threshold or distance radius (statistical) or by a minimum number of cases (combinatorial). The *size* refers to this threshold, radius, or number.

We sought to gain high-level insights into the dependency of both the performance and the calibration of localized models on cohort sizes (Collins et al. 2014). For performance, we computed the area under the receiver operating characteristic curve (AUROC) and the area under the precision–recall curve (AUPRC). Most binary classification models generate class probabilities and depend on a threshold value to classify cases; the choice of threshold ideally minimizes all kinds of classification error, and ROC and PR curves are used to visualize the trade-offs involved: The ROC curve plots the true positive rate (sensitivity) versus the true negative rate (specificity) or its complement the false positive rate, while the PR curve plots the fraction of positive-classified cases that are actually positive (precision) versus the true positive rate (recall). A greater area under either curve indicates a more accurate model, and the AUROC is much more commonly used; but the AUROC has proven to yield optimistic results when the outcome is rare (Cook and Ramadas 2020), so both are considered here as previously (Lee, Maslove, and Dubin 2015). For calibration, we computed Hosmer–Lemeshow (HL) test statistics. The HL test cannot comprehensively detect poor fit to data (Hosmer et al. 1997), but it remains a simple and useful test of overall calibration error. The distribution of the test statistic depends on the sample size, so in order for results on different units to be comparable we instead tracked its p-value. To make differences clearer, these were log-transformed. We plotted these three evaluation metrics versus size for all experiments.

To quantitatively assess reproducibility, we report the best performances using cosine similarity and combinatorial retrieval and compare both to the results of Lee, Maslove, and Dubin (2015).

To assess the effect on performance of retrieval denomination, similarity measure, and model family, we compare optimal performance (by cohort size) in folded cross-validation in a summary table of means and standard errors and in linear regression models of performance in each setting.

We compared performance under temporal validation to performance under folded cross-validation by overlaying their performance-by-size plots, with standard errors for folded cross-validation, and by calculating the difference between performances by size in units of standard errors. We note that the temporal validation training data sets (collected during CareVue) were smaller than the folded CV training sets.

### 3.5 Software

The MIMIC-III database was instantiated using the database management system PostgreSQL (PostgreSQL Global Development Group 2022). Data extraction was done through R (R Core Team 2022) using dbplyr (Wickham, Girlich, and Ruiz 2017) and RPostgres (Wickham, Ooms, and Müller 2021).

All analyses were conducted in R, using Tidyverse packages (Wickham et al. 2019). We relied additionally on proxy (Meyer and Buchta 2022). Upon acceptance for publication, our code and scripts will be made publicly available at the project repository on GitHub ^1^.

## 4 Results

Tables 1 and 2 report optimal performance measures for each batch of localized models evaluated using folded cross-validation. Figures 2 and 3 compare AUROC across care units, cohort construction method, and predictive model. Analogous plots of AUPRC and of HL test p-values are included as supplements.

**Table 1:** Optimal AUROC in 10-fold cross-validation experiments.

| MeasureModel | CCU<br>Comb | TSICU<br>Comb | CSRU<br>Comb | SICU<br>Comb | MICU<br>Comb | CCU Stat | TSICU<br>Stat | CSRU<br>Stat | SICU<br>Stat | MICU<br>Stat |
| --- | --- | --- | --- | --- | --- | --- | --- | --- | --- | --- |
| Cosine Proportion | 0.747<br>(0.045) | 0.820<br>(0.033) | 0.798<br>(0.046) | 0.773<br>(0.031) | 0.735<br>(0.019) | 0.739<br>(0.045) | 0.784<br>(0.041) | 0.754<br>(0.075) | 0.752<br>(0.022) | 0.710<br>(0.020) |
| Cosine Logistic regression | 0.752<br>(0.051) | 0.814<br>(0.035) | 0.790<br>(0.060) | 0.774<br>(0.034) | 0.736<br>(0.021) | 0.738<br>(0.053) | 0.790<br>(0.035) | 0.739<br>(0.080) | 0.758<br>(0.031) | 0.715<br>(0.022) |
| Cosine Decision tree | 0.747<br>(0.045) | 0.820<br>(0.033) | 0.798<br>(0.046) | 0.773<br>(0.031) | 0.735<br>(0.019) | 0.739<br>(0.045) | 0.784<br>(0.041) | 0.754<br>(0.075) | 0.752<br>(0.022) | 0.710<br>(0.020) |
| Gower Proportion | 0.770<br>(0.049) | 0.827<br>(0.031) | 0.856<br>(0.042) | 0.783<br>(0.029) | 0.756<br>(0.016) | 0.754<br>(0.042) | 0.798<br>(0.028) | 0.784<br>(0.066) | 0.763<br>(0.026) | 0.740<br>(0.012) |
| Gower Logistic regression | 0.768<br>(0.050) | 0.819<br>(0.039) | 0.814<br>(0.059) | 0.780<br>(0.032) | 0.754<br>(0.016) | 0.751<br>(0.051) | 0.785<br>(0.042) | 0.738<br>(0.085) | 0.757<br>(0.032) | 0.735<br>(0.015) |
| Gower Decision tree | 0.770<br>(0.049) | 0.827<br>(0.031) | 0.856<br>(0.042) | 0.783<br>(0.029) | 0.756<br>(0.016) | 0.754<br>(0.042) | 0.798<br>(0.028) | 0.784<br>(0.066) | 0.763<br>(0.026) | 0.740<br>(0.012) |

**Table 2:**
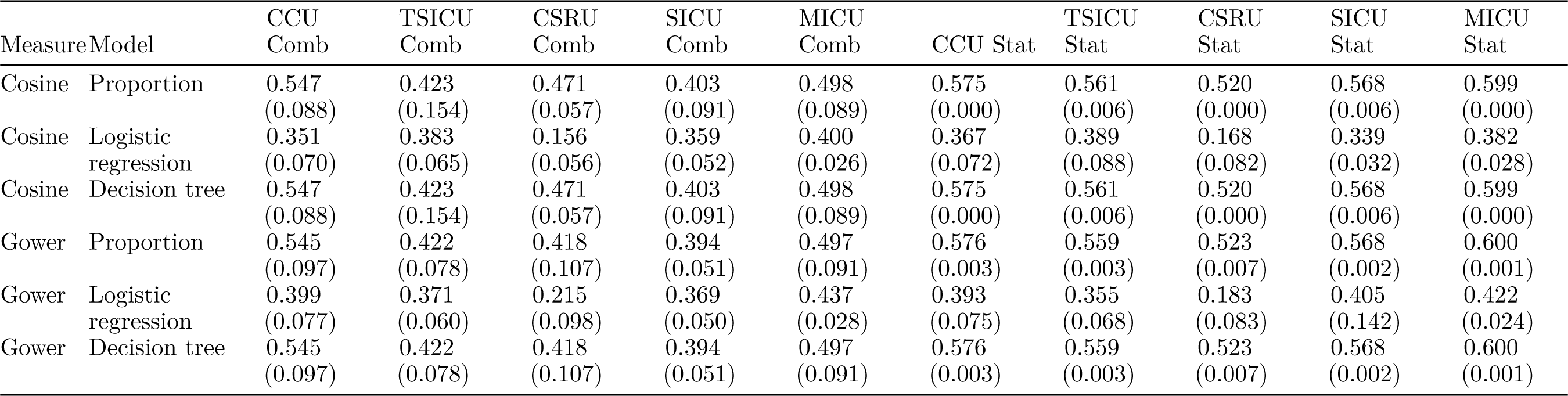
Optimal AUPRC in 10-fold cross-validation experiments.

**Figure 1:**
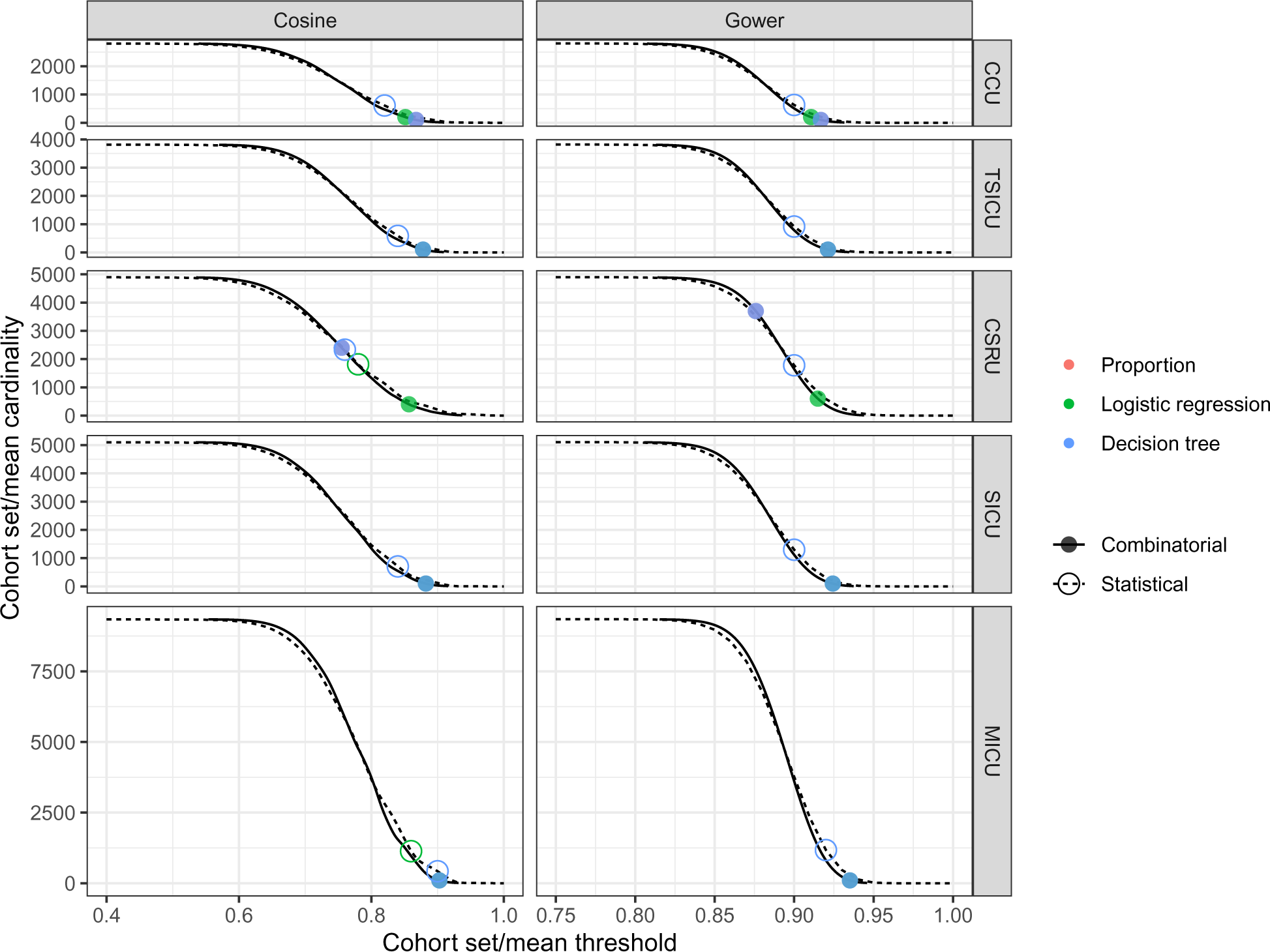
Mean similarity threshold of combinatorial cohorts and mean cardinality of statistical cohorts. Given cohorts of one denomination, the plot matches their size in that denomination to their sizes in the other denomination. AUROC-optimizing values for each model are placed on each curve.

**Figure 2:**
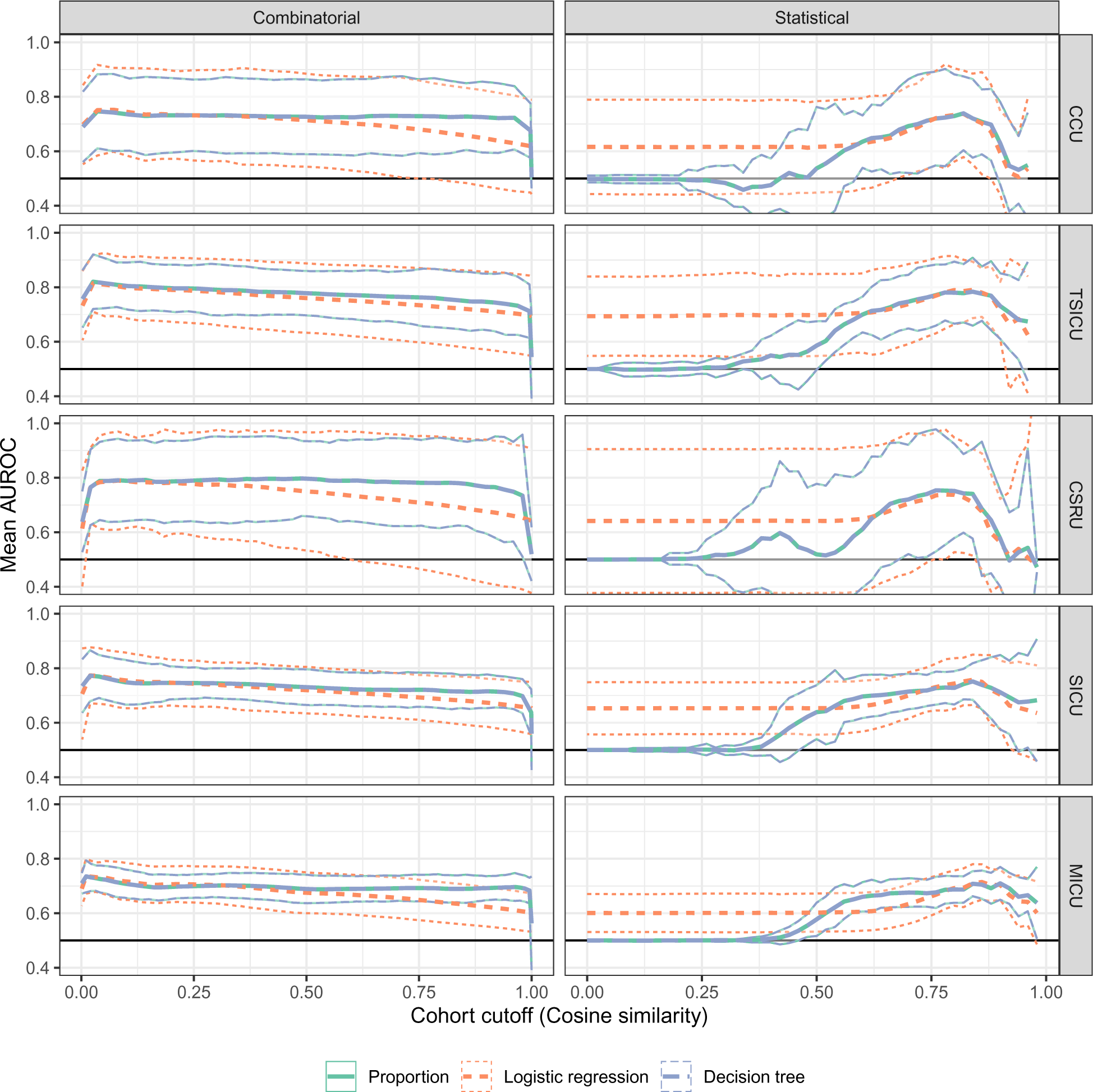
Predictive performance of localized models as cosine similarity–based cohort size parameters vary. Thick and think curves represent mean performance and 3–standard deviation confidence bands from 10-fold cross-validation.

**Figure 3:**
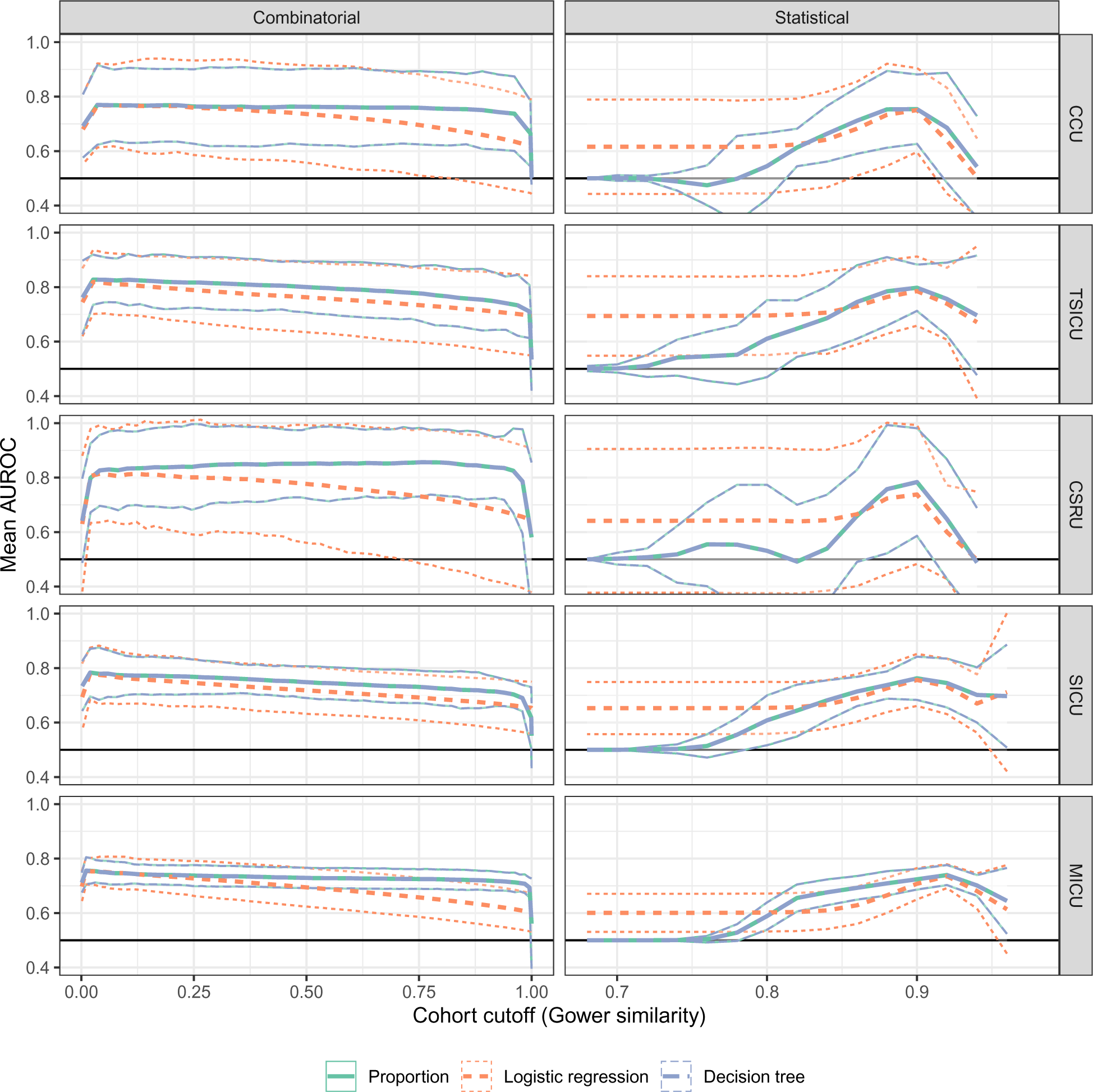
Predictive performance of localized models as Gower distance–based cohort size parameters vary. See Figure 2.

### 4.1 Reproducibility

Our results using combinatorial cohorts with cross-validation are broadly consistent with those of Lee, Maslove, and Dubin (2015). While we treat each unit separately rather than use the unit as a predictor, we obtain similar optimized mean AUROC (0.735–0.820 versus 0.753–0.830) and AUPRC (0.351–0.498, except one outlier, versus 0.347–0.474) and the relationship between each performance measure and cohort size takes a similar shape, increasing quickly to optimum performance at less than half of the training data (100–2,400 observations or 0.1–49% versus 60–6,000 observations or 0.35–35%) and slowly decreasing thereafter.

### 4.2 Choice of predictive model

While no model family consistently outperformed the others, in most cases we found that simple proportions achieved best optimal performance, followed by LR and then DT. In some instances (including MICU experiments using cosine similarity) LR models performed best, but in all CSRU experiments they performed worst. We note also that LR matched DT and often outperformed proportions with respect to AUROC when using cosine similarity; LR suffered relative to proportions and DT with respect to AUPRC.

### 4.3 Similarity measure

Localized models based on Gower distance performed only marginally better or worse than those based on cosine similarity, whether using combinatorial (AUROC 99.3–107.4%, AUPRC 88.7–137.7%) or statistical (AUROC 95.1–104.2%, AUPRC 74.7–119.5%) cohorts.

### 4.4 Cohort denomination

Combinatorial cohorts consistently produced better-performing optimized localized models than statistical cohorts.

As with combinatorial retrieval, usually less than half of the training data (4.5–58.3% for AUROC) provided the best localized predictions under statistical retrieval, arising from statistical thresholds of 0.76–0.90 (modified cosine) and 0.88–0.92 (Gower), respectively.

We note that the difference in performance between denominations defies barely detectable differences in the average sizes of the cohorts obtained. Figure 1 overlays two relationships, within the cohorts constructed for each unit: (1) As the minimum cardinality of the combinatorial cohorts changes along the abscissa, the solid curve traces the mean minimum similarity of these cohorts along the ordinate. (2) As the similarity threshold of the statistical cohorts changes along the ordinate, the dashed curve traces the mean cardinality of these cohorts along the abscissa. The sizes that optimized the AUROC under each model are marked by solid (combinatorial) and open (statistical) circles.

### 4.5 Dependency on cohort size

Notably, in all experiments DT performance as a function of cohort size closely tracked proportions performance, suggesting that the decision trees provided little performance gain. LR performance usually matched that of proportions and DT over a range of smaller cohort sizes before declining over intermediate sizes and then remaining stable over the largest cohorts. However, these three ranges depended greatly on the scale of the denomination: The intermediate range of LR underperformance dominates the combinatorial range, while the large-cohort range of LR stability and proportions/DT decline occupies most of the statistical range—even after cropping low-threshold ranges of negligible change in performance (Figure 2 and 3). This suggests that localized LR is more robust than simple proportions and DT, though at some cost to performance as cohorts grow, but perhaps more importantly that the relevant tuning range for cohort sizes is narrow for cardinality but wide for similarity threshold, relative to their theoretical ranges. (Recall that localized proportional models with combinatorial cohorts are nearest-neighbor models.)

The relative values of AUPRC were qualitatively similar to those of AUROC over the small and intermediate ranges of cohort size. However, whereas the mean AUROC of proportions and DT dropped on the largest cohorts, the mean AUPRC rose. While its variance also increased, the gain is consistent across units and similarity measures. These models thus illustrate a trade-off rather than an equivalence between AUROC and AUPRC, as theoretically discussed by J. Davis and Goadrich (2006).

Finally, the goodness of fit of the proportions, LR, and DT models, as measured by HL tests, also depended on cohort size, and this dependency varied over the same three ranges. We visualized the tests results in a supplement by their log-transformed median and IQR p-values. Calibration increased with cohort size within the small range until all models peaked. Over the medium range, LR models either were stable or improved at a slower pace, while proportion/DT models stabilized or slowly worsened. Over the largest cohorts, proportion/DT models worsened rapidly while LR models remained stable.

Taken together, these results indicate that, for most purposes, localized models are optimized while similarity cohorts are relatively small, consistent with the conclusions of Lee, Maslove, and Dubin (2015) and Lee (2017).

### 4.6 Temporal validation

In terms of the confidence bounds obtained from 10-fold CV, we find no evidence that our model families underperformed when trained on data from the CareVue database and tested on data from the MetaVision database: The AUROC and AUPRC were always within three standard errors of those of the folded-CV experiments, as illustrated using the smallest and largest units (Figure 4).

**Figure 4:**
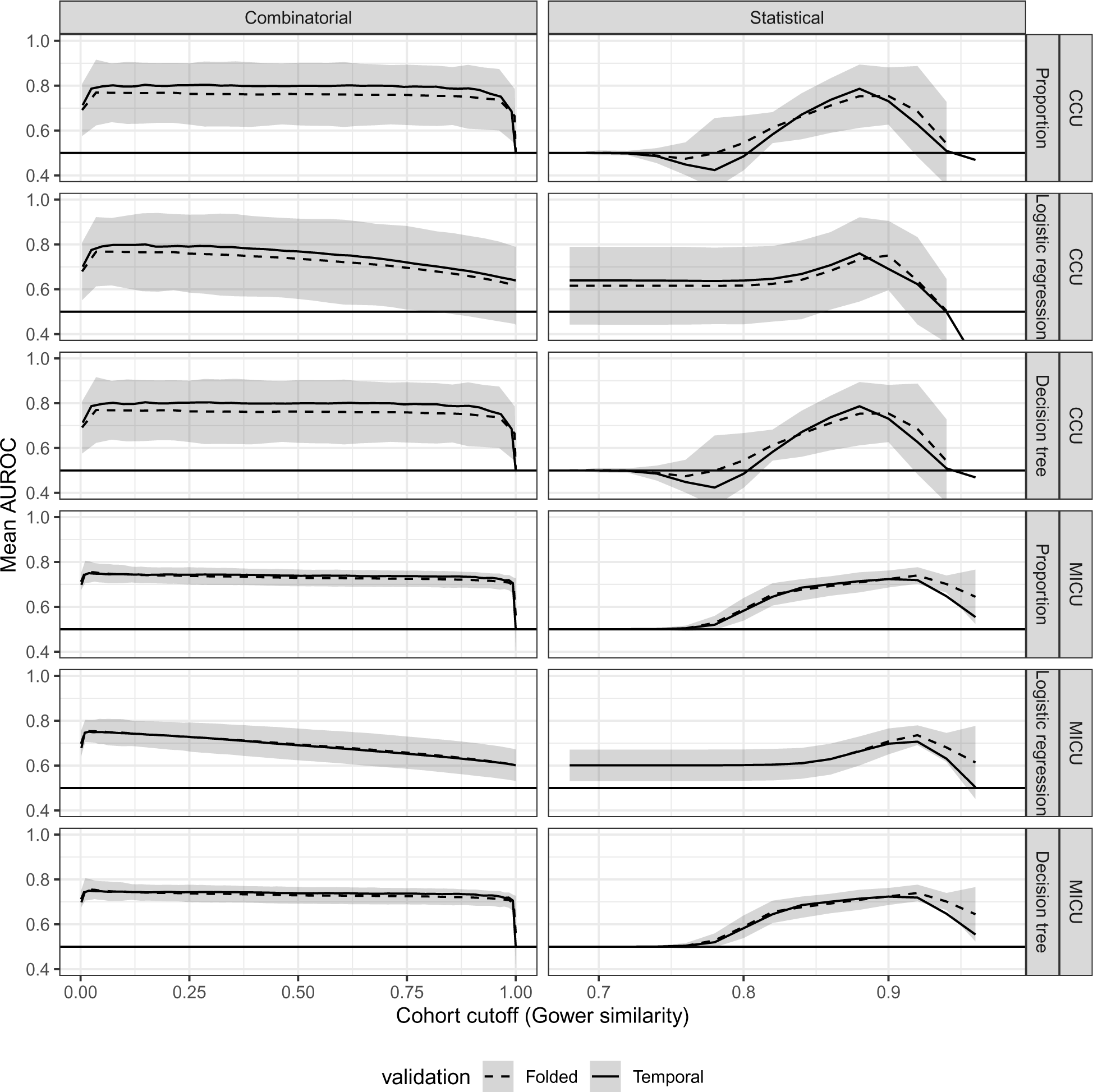
Predictive performance of localized models as Gower similarity–based cohort size parameters vary, for the smallest (CCU) and the largest (MICU) units. The shaded regions are 3–standard error confidence bands about the mean 10-fold CV performance.

**Figure 5:**
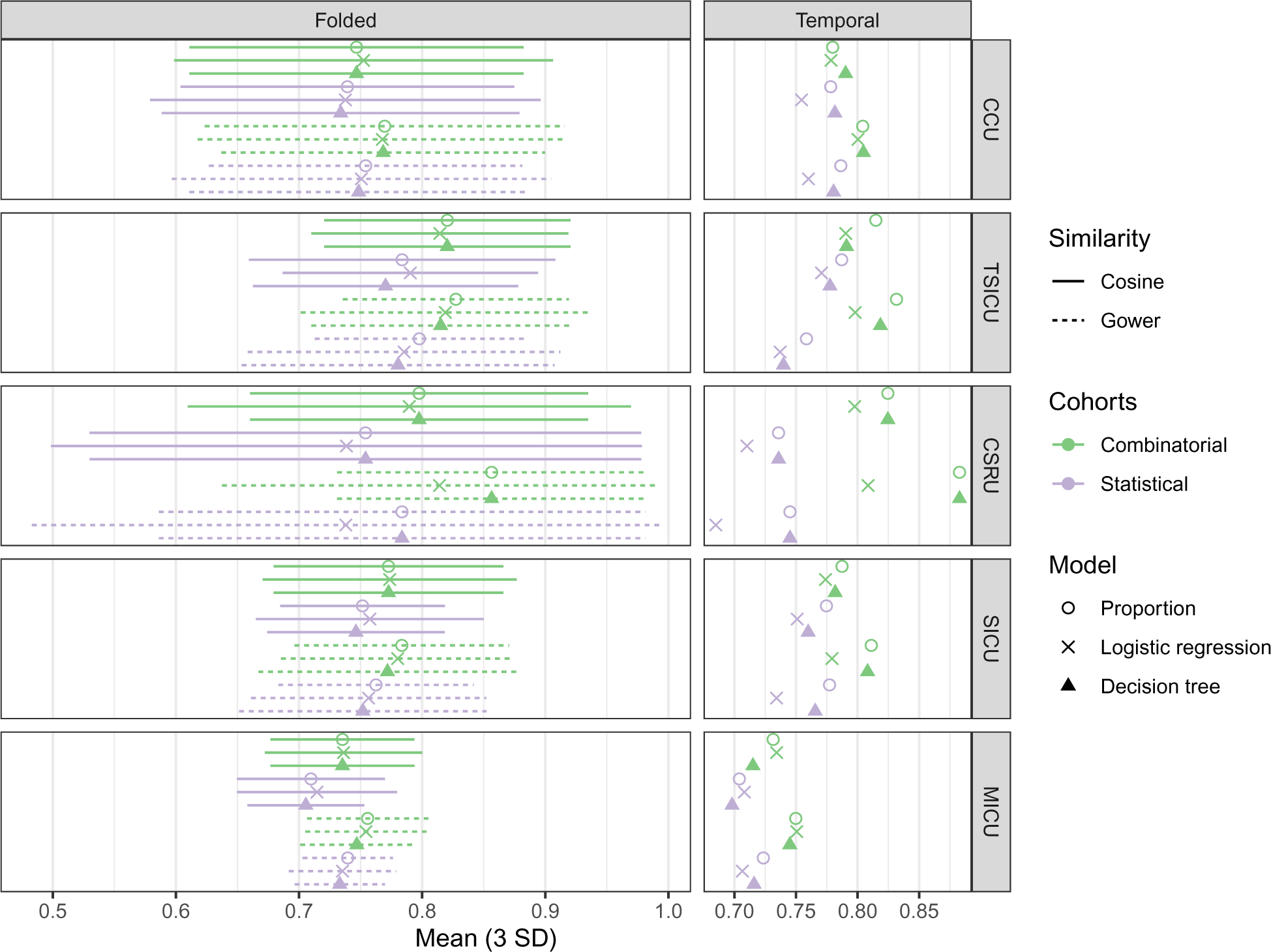

### 4.7 Relative contributions

Table 3 summarizes the estimated effects in a linear regression model of optimal AUROC on several localized modeling choices settings and choices for the folded cross-validation experiments. The largest effects are asosciated with the care unit under analysis, with the CCU as the reference value, and effects of at most a few percentage points. Well within the range of these effects is that of cohort denomination, with combinatorial cohorts achieving greater performance than statistical cohorts by 3 percentage points on average. At the lower end of these effects is that of similarity measure, with Gower similarity achieving greater performance than cosine similarity by about 1.6 percentage points. The effects associated with using the larger care units (relative to CCU) were similar in size but estimated with less confidence, while those of model choice were significantly smaller and not detectable.

**Table 3:**
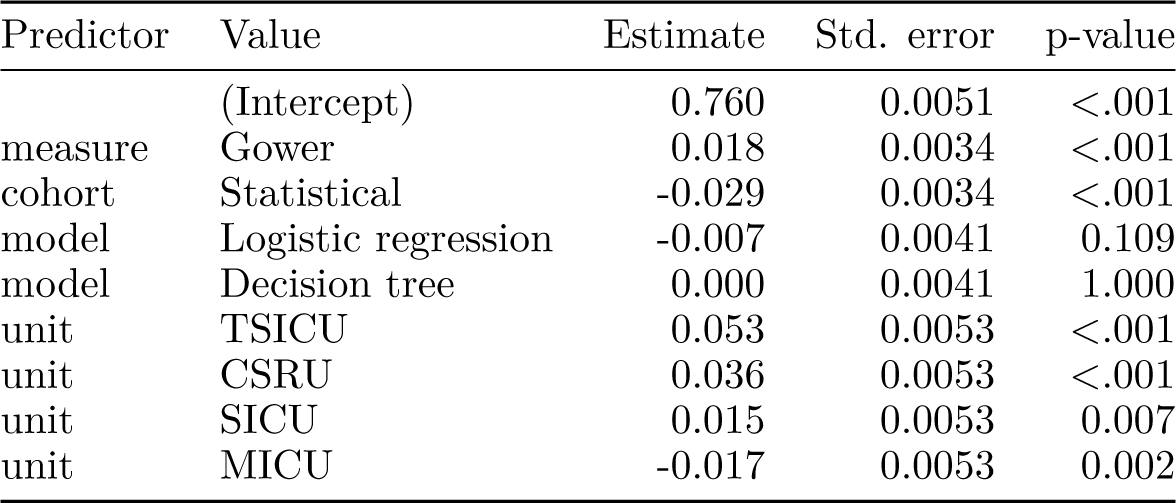
Estimated effects and standard errors in a linear model of AUROC on parameters and choices of localized models under folded cross-validation.

## 5 Discussion

We sought in this study to reproduce, robustness test, expand, and validate a novel application to mortality prediction in critical care (Lee, Maslove, and Dubin 2015) of an approach to predictive modeling that hybridizes case-based reasoning and variable-based global models. To this end, we approximately reconstituted analytic data from an updated version of the MIMIC database (Johnson, Pollard, and Mark 2016) and the previously used analysis pipeline. We then expanded this pipeline to incorporate alternative choices of similarity measure, cohort demarcation, and cross-validation. We closely reproduced previous results from scratch, and we report on the relative impact of various choices on the modeling process and on the performance of the optimized models.

Unsurprisingly but importantly, we found strong qualitative and quantitative agreement with the original study, despite separating analyses by care unit rather than pooling the critical care population. In addition to optimized performance gains, we also faithfully reproduced the shape of the relationship between localized cohort size and localized model performance observed by Lee, Maslove, and Dubin (2015) and extended this to the statistical construction. Notably, we found that the superior global performance of LR models versus using proportions (nearest neighbors prediction) and DTs is in these cases entirely an artifact of outliers: Over much of the size range, LR model performance declined while that of proportions and DT models held steady. Yet, when an amount less than 1% of the training data are excluded from localized cohorts, proportions and DTs outperformed LR; only when the least relevant cases were included did proportion and DTs underperform LR models and regress toward noninformativity.

In terms of optimized performance, we found little impact of the choice of similarity measure or validation process. Regarding the similarity measure, we caution that results still depend greatly on the choice of predictors. For example, in unreported results, we found much worse performance using the modified cosine similarity on an analytic data set comprising only demographic variables and frequent itemsets of diagnosis codes in patients’ medical history.

Regarding validation, serious challenges have been documented to the real-world application of EHR-based predictive models, among them the risk of performance drift as models are evaluated using new cases progressively far into the future (S. E. Davis et al. 2018) and the need for temporal validation to better simulate performance on new cases in the near future (Major, Jethani, and Aphinyanaphongs 2020). In this study, we did not observe weaker performance on temporally-partitioned holdout data relative to folded cross-validation.^2^ Because the data were collected and stored for research use by the same team, this amount of agreement may not be surprising, but it does raise the possibility that some temporal invalidations may be due more to differences in the collection and processing of data than to changes in the real-world presentation and management of disease.

The denomination of localized cohorts—statistical versus combinatorial—mattered little to optimized performance, despite its dramatic effect on how the relationship between cohort size and performance is presented to and understood by the investigator. This result challenges the conclusions of Park, Kim, and Chun (2006) that statistical retrieval yields better predictive performance than combinatorial retrieval in classical CBR. We also observed a tight relationship between the prescribed size of localized cohorts of one denomination—the minimum cardinalities of combinatorial cohorts and the thresholds of statistical cohorts—and the average sizes according to the other. The lack of a clear advantage between the denominations may be attributable to this close average relationship. Whether this average is representative, and which cases, if any, nevertheless receive less reliable predictions using one denomination versus the other, is a topic for future study.

Our most important result may be that the impact of the adaptation method (the choice of predictive model) was smaller than that of the retrieval method (the similarity measure and the cohort denomination) by a factor of between 2 and 6. The lesser impact of adaptation is suggestive of the “performance plateau” that may be achieved by a large variety of predictive models even as a subset of them reliably produce marginally better performance (Hand 2006); in contrast, retrieval is the means by which localized models choose their study populations, so the still-small impact of this choice could be surprising in light of the reported performance gains of traditional case-based reasoning (Sharafoddini, Dubin, and Lee 2017).

### 5.1 Limitations

While our objective is to evaluate the reproducibility and robustness of localized models, this study does not provide direct insight into their real-world performance. In particular, the data we used are highly curated and not necessarily representative of critical care of other EHR-derived data “in the wild”. Moreover, AUROC and AUPRC, while useful summary statistics for binary outcome models, are not primary clinical considerations for predictive models, which depend on clinical settings and on patient and physician goals to be calibrated and interpreted. The choices and parameters we considered represent only a fraction of the diversity of localized and similar modeling approaches that have been proposed, many of which are highly tailored to their specific use cases, as discussed in the forthcoming review.

### 5.2 Future work

The validation and reproduction of models built on health data is notoriously difficult but has been made much more achievable by several developments in data availability and standardization and in evaluation techniques. In addition to the release of larger and richer health data sets, the rise of standards such as SNOMED, PCORNet, and FHIR have helped make data collected and stored at different institutions available for research that follows fixed protocols, including reproduction, validation, and extension of past studies, though still many additional uncontrolled factors remain. Temporal validation has been shown to better predict external validation results and stands to inform and address key practical problems such as performance drift (Major, Jethani, and Aphinyanaphongs 2020).

Based on our approach, statistical cohorts are easier to fine-tune than combinatorial cohorts, in that localized models are optimized within a narrower relative range of cardinality than of similarity. This presents a trade-off against the slightly stronger performance of combinatorial cohorts, though the averages sizes of cohorts of both denominations track closely. The significance, or not, of how their sizes are distributed may be an important question for the value of localized models for outlying cases.

Several reproducibility considerations were not relevant to or without the scope of this study but deserve mention. The proliferation of data and the rise of standards herald new problems, including differences in meaning and completeness arising from differences in practice. Methods to account for these patterns while maintaining both performance and interpretability are nascent and will need continued attention (Nijman et al. 2022). The prospects of ensemble models are good for predictive performance (Mahajan et al. 2023), and this technique could greatly benefit a localized approach, in which the bulk of the computational cost is incurred by cohort construction. The prospects for interpreting such localized ensembles would need to be determined for this hybrid approach to prove useful.

### 5.3 Conclusions

In summary, we qualitatively and quantitatively reproduced previously reported success in predicting 30-day mortality despite an expanded study population, restriction of experiments to individual units, and several technical challenges to a faithful reproduction of the methods. While results varied significantly by population, few methodological choices noticeably impacted the optimized model performance; most importantly and surprisingly, parameters governing cohort retrieval had larger and more evident effects than the more commonly tested choice of predictive model family. Our results reinforce the conclusions of Lee, Maslove, and Dubin (2015) and Lee (2017) that localized models *can* outperform global models but also challenge their findings that the choice of model is highly consequential. Instead, our results suggest that these localized models perform little better than routine nearest neighbors prediction (there called “death count”), and in some cases worse. Given that studies of localized modeling urge evaluation in diverse settings yet do not consistently report improved performance over common alternatives, the approach faces an acute need for such comparisons.

## Data Availability

All data used in the present study are available online at PhysioNet. All code used to conduct the analysis will be published upon acceptance for journal publication and will be made available on request in the meantime.

https://physionet.org/content/mimiciii/1.4/

## 6 Acknowledgments

Pre-processing and analysis scripts built upon previous work on MIMIC-III by Lauren Nicole Geiser and Beverly Anne Setzer. Henrique de Assis Lopes Ribeiro provided valuable comments on a draft of this manuscript.

## Supplement

Figures 6 and 7 compare AUPRC, and Figures 8 and 9 compare log-transformed HL test p-values, across care units, cohort construction method, and predictive model.

**Figure 6:**
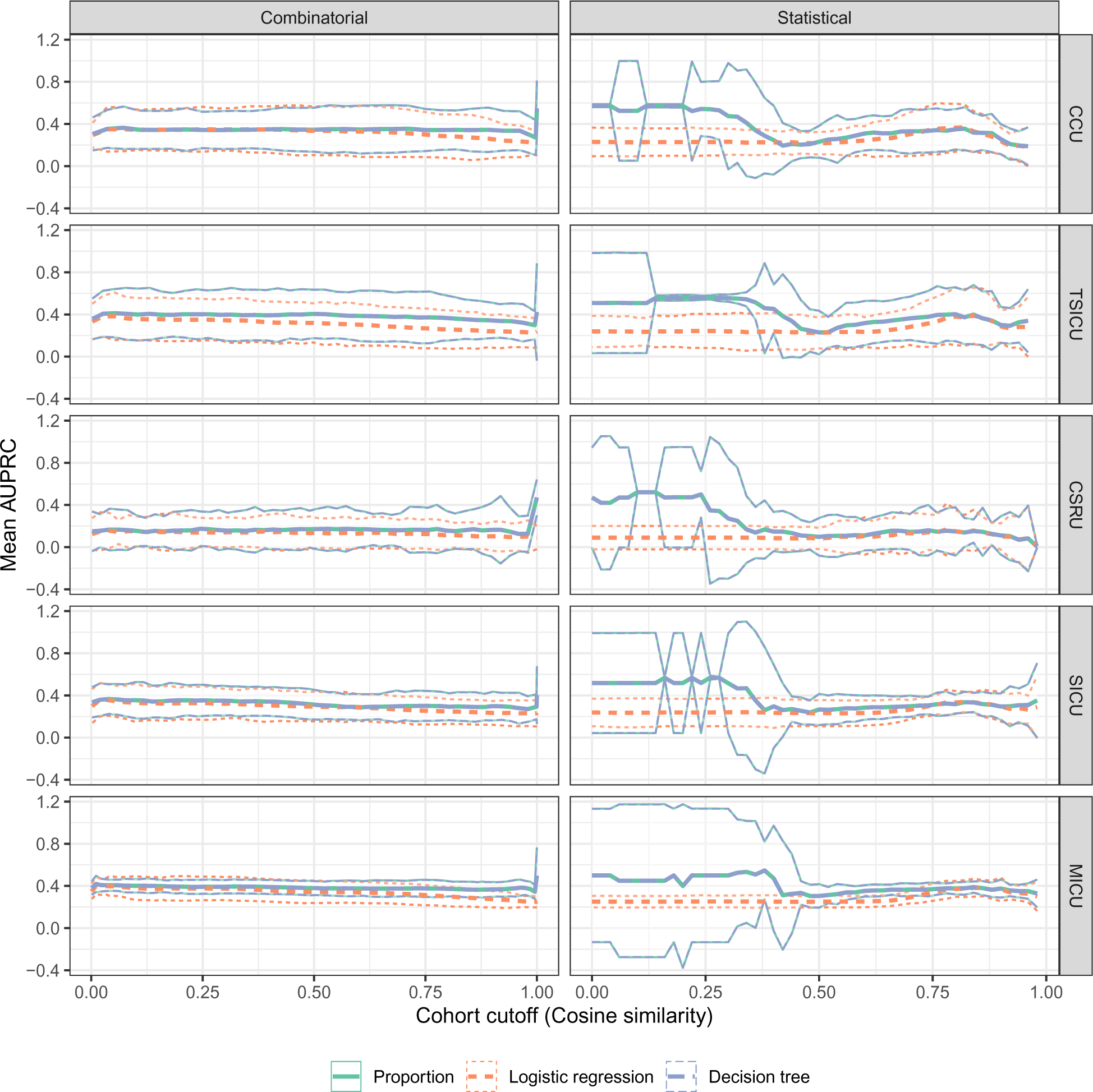
Predictive performance of localized models as cosine similarity–based cohort size parameters vary. Thick and think curves represent mean performance and 3–standard deviation confidence bands from 10-fold cross-validation.

**Figure 7:**
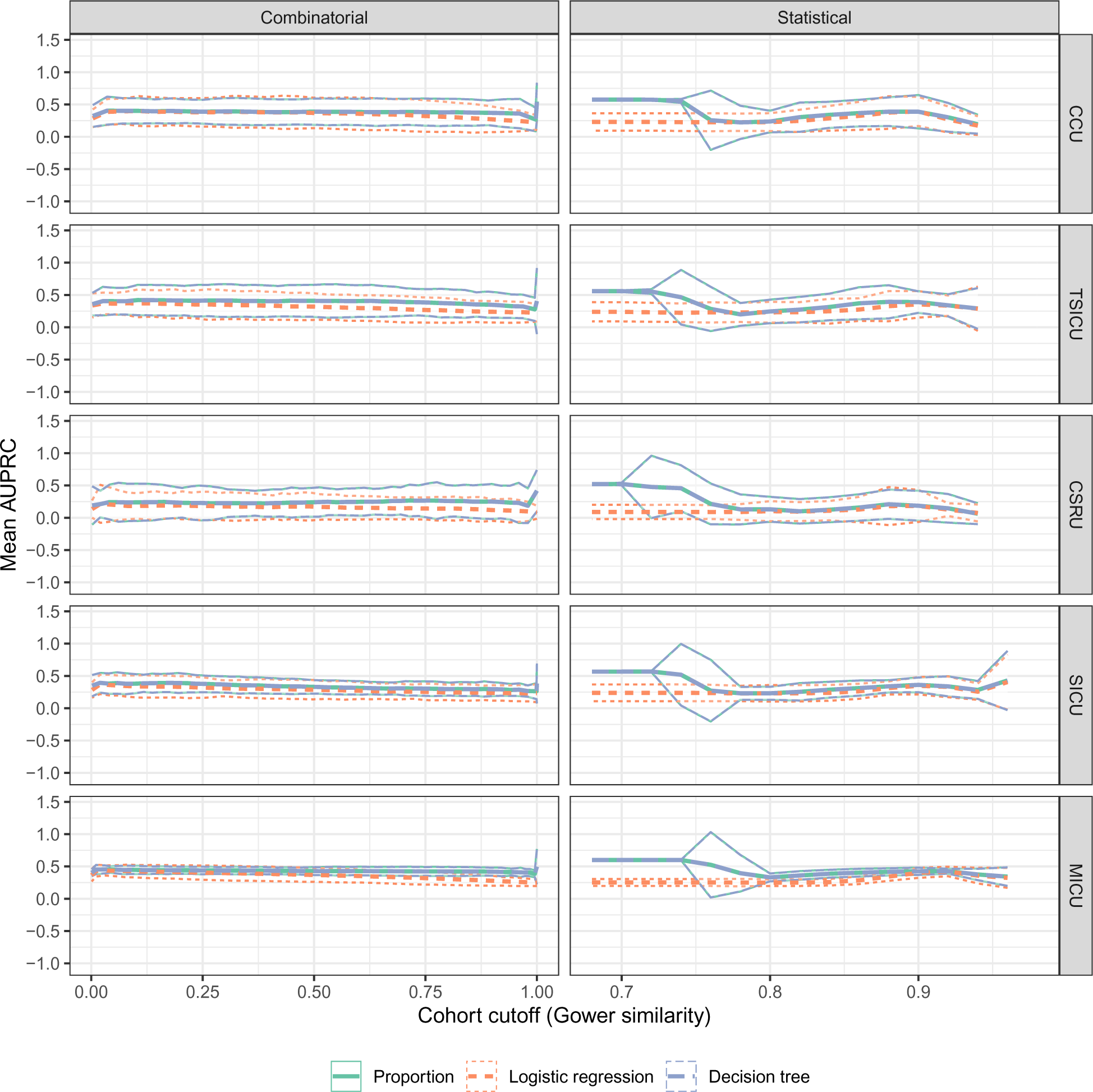
Predictive performance of localized models as Gower distance–based cohort size parameters vary. See Figure 6.

**Figure 8:**
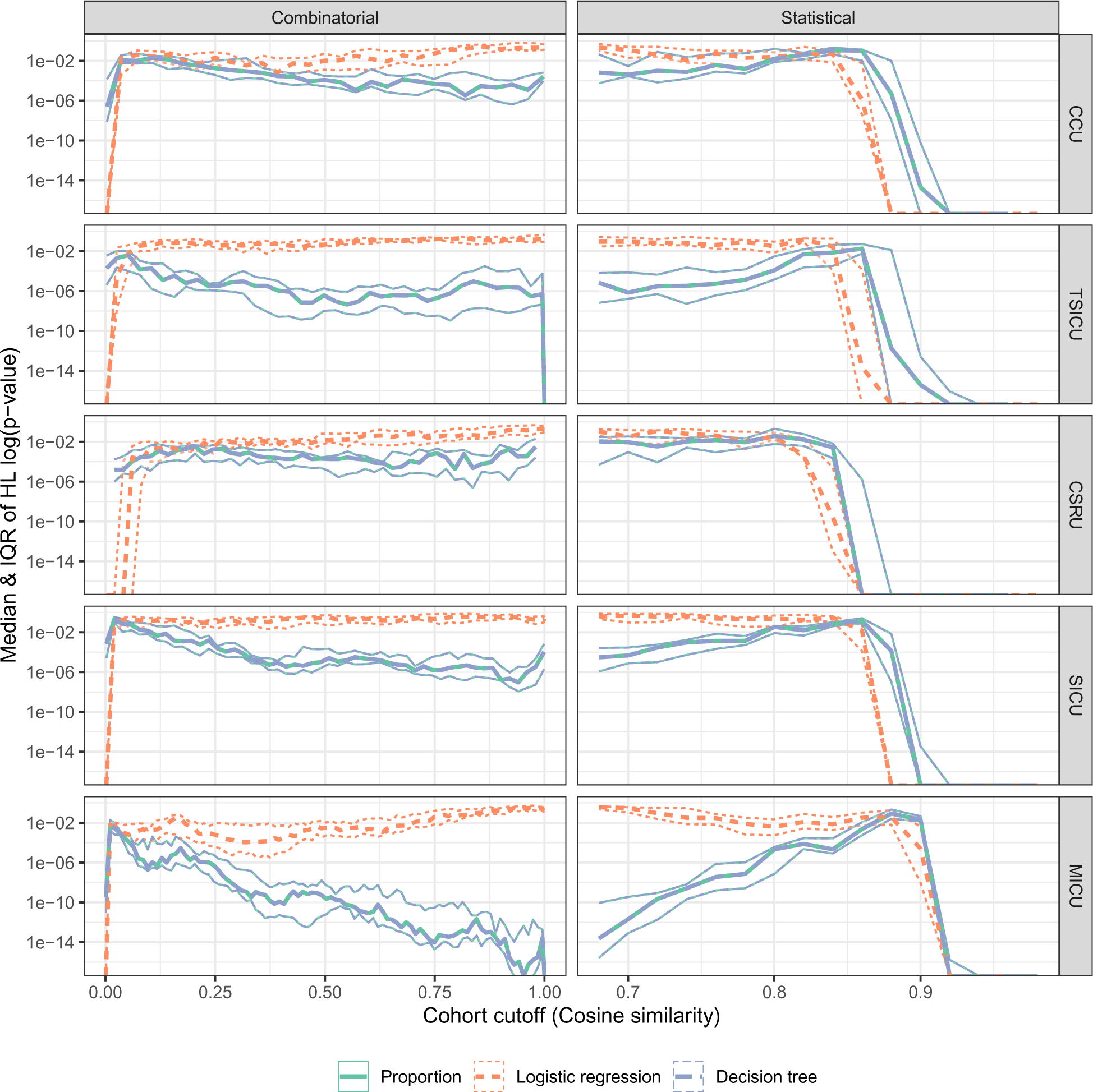
Goodness of fit of localized models as cosine similarity–based cohort size parameters vary. Thick and think curves represent mean performance and 3–standard deviation confidence bands from 10-fold cross-validation.

**Figure 9:**
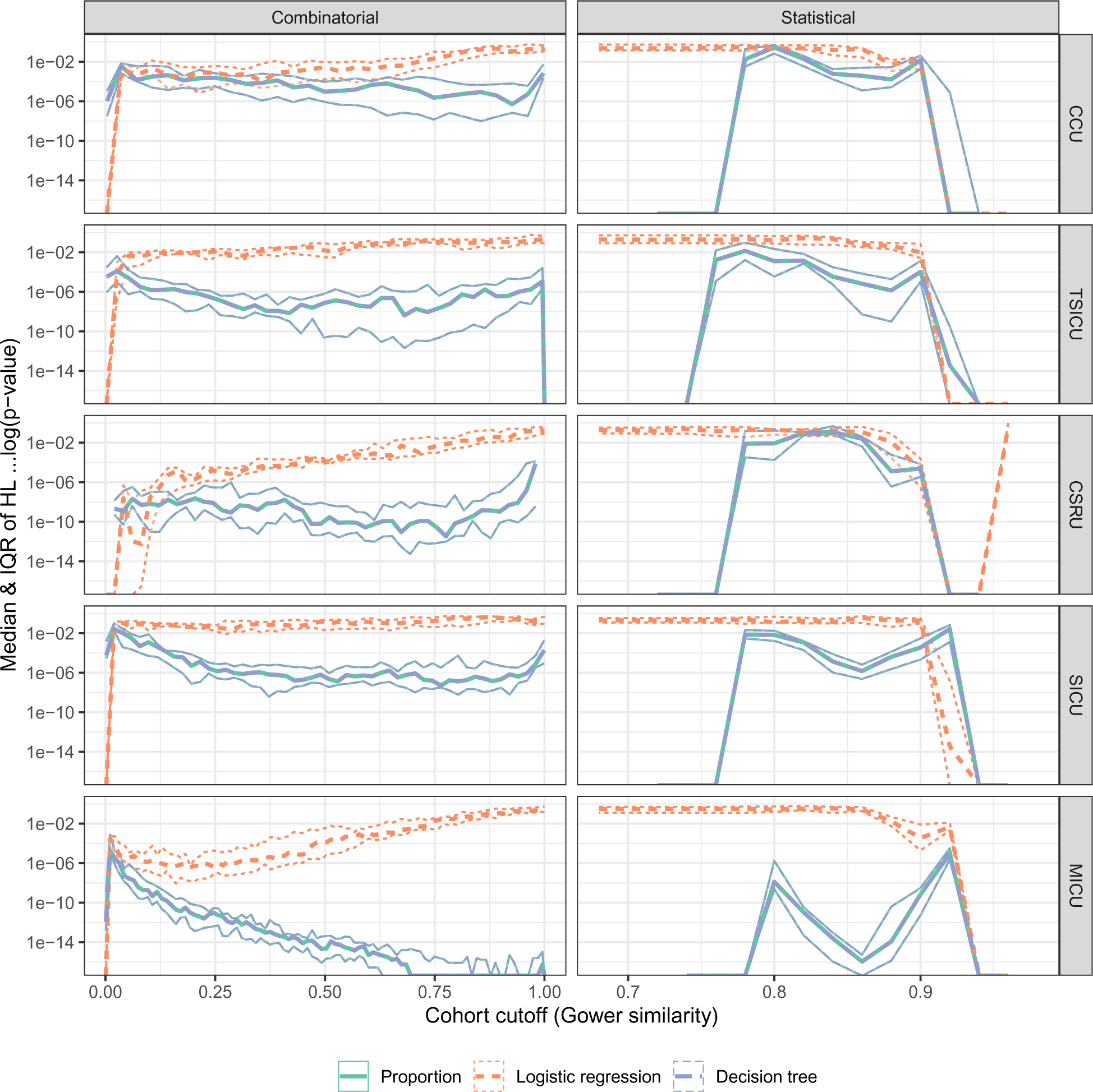
Goodness of fit of localized models as Gower distance–based cohort size parameters vary. See Figure 8.

## Footnotes

1 https://github.com/amy-work/UCONN-Health-project

2 I think we should account for partition size by comparing the temporal validation results to validation with (say, 10) random partitions of the same size.

## Notes

### Competing Interest Statement

The authors have declared no competing interest.

### Funding Statement

This study did not receive specific funding.

### Author Declarations

The study used only openly available human data derived from electronic health records that are maintained and can be accessed at PhysioNet upon completion of a short course: https://physionet.org/content/mimiciii/1.4/

## References

Bichindaritz, Isabelle, and Cindy Marling. 2006. “Case-Based Reasoning in the Health Sciences: What’s Next?” Artificial Intelligence in Medicine, Case-based reasoning in the health sciences, 36 (2): 127–35. 10.1016/j.artmed.2005.10.008.

Bichindaritz, Isabelle, and Stefania Montani. 2011. “Advances in Case-Based Reasoning in the Health Sciences.” Artificial Intelligence in Medicine, Advances in Case-Based Reasoning in the Health Sciences, 51 (2): 75–79. 10.1016/j.artmed.2011.01.001.

Collins, Gary S., Joris A. de Groot, Susan Dutton, Omar Omar, Milensu Shanyinde, Abdelouahid Tajar, Merryn Voysey, et al. 2014. “External Validation of Multivariable Prediction Models: A Systematic Review of Methodological Conduct and Reporting.” BMC Medical Research Methodology 14 (1): 40. 10.1186/1471-2288-14-40.

Cook, Jonathan, and Vikram Ramadas. 2020. “When to Consult Precision-Recall Curves.” The Stata Journal 20 (1): 131–48. 10.1177/1536867X20909693.

Davis, Jesse, and Mark Goadrich. 2006. “The Relationship Between Precision-Recall and ROC Curves.” In Proceedings of the 23rd International Conference on Machine Learning, 233–40. ICML ’06. New York, NY, USA: Association for Computing Machinery. 10.1145/1143844.1143874.

Davis, Sharon E., Thomas A. Lasko, Guanhua Chen, and Michael E. Matheny. 2018. “Calibration Drift Among Regression and Machine Learning Models for Hospital Mortality.” AMIA Annual Symposium Proceedings 2017 (April): 625–34.

Goldberger, Ary L., Luis A. N. Amaral, Leon Glass, Jeffrey M. Hausdorff, Plamen Ch. Ivanov, Roger G. Mark, Joseph E. Mietus, George B. Moody, Chung-Kang Peng, and H. Eugene Stanley. 2000. “PhysioBank, PhysioToolkit, and PhysioNet: Components of a New Research Resource for Complex Physiologic Signals.” Circulation 101 (23): e215–20. 10.1161/01.CIR.101.23.e215.

Goyal, Navin, Yury Lifshits, and Hinrich Schütze. 2008. “Disorder Inequality: A Combinatorial Approach to Nearest Neighbor Search.” In Proceedings of the 2008 International Conference on Web Search and Data Mining, 25–32. WSDM ’08. New York, NY, USA: Association for Computing Machinery. 10.1145/1341531.1341538.

Hand, David J. 2006. “Classifier Technology and the Illusion of Progress.” Statistical Science 21 (1): 1–14. 10.1214/088342306000000060.

Hosmer, D. W., T. Hosmer, S. Le Cessie, and S. Lemeshow. 1997. “A Comparison of Goodness-of-Fit Tests for the Logistic Regression Model.” Statistics in Medicine 16 (9): 965–80. 10.1002/(SICI)1097-0258(19970515)16:9%3C965::AID-SIM509%3E3.0.CO;2-O.

Johnson, Alistair E. W., Tom J. Pollard, and Roger G. Mark. 2016. “MIMIC-III Clinical Database.” PhysioNet. 10.13026/C2XW26.

Johnson, Alistair E. W., Tom J. Pollard, Lu Shen, Li-wei H. Lehman, Mengling Feng, Mohammad Ghassemi, Benjamin Moody, Peter Szolovits, Leo Anthony Celi, and Roger G. Mark. 2016. “MIMIC-III, a Freely Accessible Critical Care Database.” Scientific Data 3 (1). 10.1038/sdata.2016.35.

Kasabov, Nikola, and Yingjie Hu. 2010. “Integrated Optimisation Method for Personalised Modelling and Case Studies for Medical Decision Support.” International Journal of Functional Informatics and Personalised Medicine 3 (3): 236–56. 10.1504/IJFIPM.2010.039123.

Lee, Joon. 2017. “Patient-Specific Predictive Modeling Using Random Forests: An Observational Study for the Critically Ill.” JMIR Medical Informatics 5 (1): e3. 10.2196/medinform.6690.

Lee, Joon, David M. Maslove, and Joel A. Dubin. 2015. “Personalized Mortality Prediction Driven by Electronic Medical Data and a Patient Similarity Metric.” Edited by Frank Emmert-Streib. PLOS ONE 10 (5): e0127428. 10.1371/journal.pone.0127428.

Liang, W, YJ Hu, and N Kasabov. 2015. “Evolving Personalized Modeling System for Integrated Feature, Neighborhood and Parameter Optimization Utilizing Gravitational Search Algorithm.” Evolving Systems 6 (1): 1–14. 10.1007/s12530-013-9081-x.

Lowsky, D. J., Y. Ding, D. K. K. Lee, C. E. McCulloch, L. F. Ross, J. R. Thistlethwaite, and S. A. Zenios. 2013. “A K-nearest Neighbors Survival Probability Prediction Method.” Statistics in Medicine 32 (12): 2062–69. 10.1002/sim.5673.

Ma, X, YB Si, ZF Wang, and YQ Wang. 2020. “Length of Stay Prediction for ICU Patients Using Individualized Single Classification Algorithm.” Computer Methods and Programs in Biomedicine 186. 10.1016/j.cmpb.2019.105224.

Mahajan, Palak, Shahadat Uddin, Farshid Hajati, and Mohammad Ali Moni. 2023. “Ensemble Learning for Disease Prediction: A Review.” Healthcare 11 (12): 1808. 10.3390/healthcare11121808.

Major, Vincent J, Neil Jethani, and Yindalon Aphinyanaphongs. 2020. “Estimating Real-World Performance of a Predictive Model: A Case-Study in Predicting Mortality.” JAMIA Open 3 (2): 243–51. 10.1093/jamiaopen/ooaa008.

Mariuzzi, G., A. Mombello, L. Mariuzzi, P. W. Hamilton, J. E. Weber, D. Thompson, and P. H. Bartels. 1997. “Quantitative Study of Ductal Breast Cancer–Patient Targeted Prognosis: An Exploration of Case Base Reasoning.” Pathology - Research and Practice 193 (8): 535–42. 10.1016/S0344-0338(97)80011-8.

Meyer, David, and Christian Buchta. 2022. “Proxy: Distance and Similarity Measures.”

Ng, Kenney, Jimeng Sun, Jianying Hu, and Fei Wang. 2015. “Personalized Predictive Modeling and Risk Factor Identification Using Patient Similarity.” AMIA Joint Summits on Translational Science Proceedings. AMIA Joint Summits on Translational Science 2015: 132–36.

Ng, K, U Kartoun, H Stavropoulos, JA Zambrano, and PC Tang. 2021. “Personalized Treatment Options for Chronic Diseases Using Precision Cohort Analytics.” Scientific Reports 11 (1). 10.1038/s41598-021-80967-5.

Nijman, S. W. J., A. M. Leeuwenberg, I. Beekers, I. Verkouter, J. J. L. Jacobs, M. L. Bots, F. W. Asselbergs, K. G. M. Moons, and T. P. A. Debray. 2022. “Missing Data Is Poorly Handled and Reported in Prediction Model Studies Using Machine Learning: A Literature Review.” Journal of Clinical Epidemiology 142 (February): 218–29. 10.1016/j.jclinepi.2021.11.023.

Park, Yoon-Joo, Byung-Chun Kim, and Se-Hak Chun. 2006. “New Knowledge Extraction Technique Using Probability for Case-Based Reasoning: Application to Medical Diagnosis.” Expert Systems 23 (1): 2–20. 10.1111/j.1468-0394.2006.00321.x.

PostgreSQL Global Development Group. 2022. “PostgreSQL.”

R Core Team. 2022. “R: A Language and Environment for Statistical Computing.” Vienna, Austria: R Foundation for Statistical Computing.

Sharafoddini, Anis, Joel A Dubin, and Joon Lee. 2017. “Patient Similarity in Prediction Models Based on Health Data: A Scoping Review.” JMIR Medical Informatics 5 (1): e7. 10.2196/medinform.6730.

Verma, A, M Fiasche, M Cuzzola, and G Irrera. 2015. “Comparative Study of Existing Personalized Approaches for Identifying Important Gene Markers and for Risk Estimation in Type2 Diabetes in Italian Population.” Evolving Systems 6 (1): 15–22. 10.1007/s12530-013-9083-8.

Vilhena, João, Henrique Vicente, M. Rosário Martins, José M. Grañeda, Filomena Caldeira, Rodrigo Gusmão, João Neves, and José Neves. 2016. “A Case-Based Reasoning View of Thrombophilia Risk.” Journal of Biomedical Informatics 62: 265–75. 10.1016/j.jbi.2016.07.013.

Wang, Ni, Yanqun Huang, Honglei Liu, Xiaolu Fei, Lan Wei, Xiangkun Zhao, and Hui Chen. 2019. “Measurement and Application of Patient Similarity in Personalized Predictive Modeling Based on Electronic Medical Records.” BioMedical Engineering OnLine 18 (1). 10.1186/s12938-019-0718-2.

Wang, Yuning, Yingzi Liang, Hui Sun, and Yufei Yang. 2020. “Emergency Response for COVID-19 Prevention and Control in Urban Rail Transit Based on Case-Based Reasoning Method.” Discrete Dynamics in Nature & Society, December, 1–9.

Wickham, Hadley, Mara Averick, Jennifer Bryan, Winston Chang, Lucy D’Agostino McGowan, Romain François, Garrett Grolemund, et al. 2019. “Welcome to the Tidyverse.” Journal of Open Source Software 4 (43): 1686. 10.21105/joss.01686.

Wickham, Hadley, Maximilian Girlich, and Edgar Ruiz. 2017. “Dbplyr: A ’Dplyr’ Back End for Databases.”

Wickham, Hadley, Jeroen Ooms, and Kirill Müller. 2021. “RPostgres: Rcpp Interface to PostgreSQL.”

